# Oral Swab Xpert MTB/RIF Ultra for Tuberculosis Diagnosis in Ethiopian Prisons

**DOI:** 10.64898/2026.03.07.26347730

**Authors:** Kelemework Adane, Russell R Kempker, Kidist Bobosha, Abyot Meaza, Henry M Blumberg

**Author notes:** Correspondence: Kelemework Adane.

## Abstract

As asymptomatic (subclinical) tuberculosis (TB) is increasingly recognized, there is a need for sputum-free diagnostic approaches applicable to both symptomatic individuals and population screening. Oral swab analysis (OSA) using Xpert MTB/RIF Ultra (Ultra) is a promising approach; however, its performance has not been evaluated in prisons. We conducted a cross-sectional study to assess the diagnostic accuracy of oral swab Ultra for TB diagnosis in Ethiopian prisons. Incarcerated adults (≥18 years) presenting with a cough of any duration were enrolled. Oral swabs were collected and processed using a customized 2:1 sample-to-reagent protocol and tested with Ultra. Diagnostic accuracy was evaluated against a composite microbiological reference standard (MRS) comprising sputum Ultra and liquid culture. Among 858 incarcerated men screened across two prisons and one detention center, 240 were eligible, and 221 were included in the analysis. Overall, 34 participants (15.4%) were diagnosed with pulmonary TB using MRS. Compared with the composite MRS, oral swab Ultra detected 21 cases, yielding a sensitivity of 61.8% (95% CI, 44.7–76.6%) and a specificity of 100%. Sensitivity compared with sputum Ultra was 63.6% (95% CI, 45.1–79.6%), with 100% specificity. Performance varied by bacillary burden: sensitivity reached 100% among participants with medium or high sputum bacillary loads, whereas none of the seven cases with trace bacillary loads were detected using oral swabs. In summary, oral swab Xpert Ultra, using a customized processing protocol, demonstrated moderate-to-high sensitivity for TB diagnosis in high-burden Ethiopian prisons, supporting its potential role as a complementary tool in prison TB screening initiatives.

**IMPORTANCE:** Prisons in high-burden countries such as Ethiopia remain heavily affected by undiagnosed tuberculosis (TB). Overreliance on sputum samples for diagnosis is a key contributing factor. Sputum-based tests may fail to capture all TB cases in such settings, as many incarcerated individuals with active TB are either unable to produce sputum or have asymptomatic (subclinical) disease. Patients with minimal or no symptoms (subclinical or asymptomatic TB), on the other hand, contribute to ongoing transmission. Oral swab analysis (OSA) using molecular diagnostics such as Xpert MTB/RIF Ultra (Ultra) offers a promising, non-invasive option for TB diagnosis. However, OSA with Ultra has not been systematically evaluated for TB screening in prison settings in high-burden regions, where diagnostic needs are greatest. In this study, we show that oral swab testing with Xpert Ultra is feasible and demonstrates moderate-to-high sensitivity for TB diagnosis in Ethiopian prisons.

## INTRODUCTION

Tuberculosis (TB), caused by *Mycobacterium tuberculosis* (Mtb), remains a major global public health threat and is the world’s deadliest disease from a single etiologic agent (1). Incarcerated individuals are disproportionately at high risk of acquiring and developing TB. The pooled TB prevalence in prisons in high-burden countries is 3.5 which is over 30 times higher than in the general population and even higher in sub-Saharan Africa (SSA) (7.7%) (2–4). Ethiopia bears a particularly high burden of TB, with a pooled prevalence of 9.8% in its prisons (5). Prisons can amplify the transmission of Mtb, increasing the risk of spillover to surrounding communities through inmates, staff, and visitors (6, 7). Early TB screening and treatment in prisons are essential to interrupt transmission and enhance clinical outcomes.

Diagnosis is the weakest link in the TB cascade of care, with millions of TB cases going undiagnosed annually (1, 8). Such diagnostic gaps are especially evident in prisons in low- and middle-income countries (LMICs), where diagnostic capacity is frequently inadequate (9, 10). Overreliance on sputum-based testing is a key contributor to the undiagnosed cases, as many people with TB may not produce sputum and/or are asymptomatic (11). Patients with minimal or no symptoms (subclinical or asymptomatic TB) also contribute to ongoing transmission (12–14). Moreover, the lack of basic infrastructure for sputum collection, including designated spaces for the segregation of symptomatic individuals, further worsens the problem.

The World Health Organization (WHO) has prioritized non-invasive, sputum-free diagnostic methods and systematic TB screening in incarcerated individuals (15). Oral swab analysis (OSA) using Xpert MTB/RIF Ultra (Ultra; Cepheid, Sunnyvale, CA, USA) offers a promising, non-invasive option for TB diagnosis (16). Research employing OSA with Ultra testing has demonstrated variable sensitivity among symptomatic adults, ranging from 45.0% to 78.1% compared to the reference standard of sputum Ultra and/or culture (17–22). However, OSA with Ultra has not been systematically evaluated for TB screening in prison settings in SSA, where diagnostic needs are greatest.

One of the major challenges in OSA is technical challenges with sample processing, which can affect diagnostic sensitivity (17). The Xpert Sample Reagent (SR; Cepheid, USA), which employs a 2:1 SR-to-sample ratio (resulting in a final SR concentration of 66.7%), was optimized for use with sputum (23). Applying the same protocol to oral swab samples may result in over-dilution and reduced DNA recovery, particularly given the typically low bacillary load in oral specimens. Several studies using existing sputum-based Ultra protocol have shown that SR can effectively process oral swabs; however, diagnostic sensitivity was variable and often suboptimal (17, 24). Customized oral swab processing approaches that use lower volumes of SR may improve the diagnostic sensitivity of oral swab Ultra and increase its reliability in real-world screening programs.

In an analytical study conducted by Chilambi and colleagues from the Foundation for Innovative New Diagnostics (FIND), it was demonstrated that reducing the final concentration of the SR in tongue swab processing for the Ultra substantially improves the analytical sensitivity (24). However, evidence on the performance of this modified approach in real-world, high-burden settings remains limited. To further address, we evaluated the diagnostic accuracy and operational feasibility of oral swab–based Xpert Ultra using this customized sample processing protocol for TB detection in Ethiopian prison settings.

## MATERIALS AND METHODS

### Ethics Statement

Ethical approval was obtained from the Institutional Research Ethics Review Committee (IRERC) of the College of Health Sciences of Addis Ababa University (approval number: AAUMF03/008). Written informed consent was obtained from all participants. Participant confidentiality was ensured using unique IDs and password-protected files.

### Study Design and Setting

This cross-sectional study enrolled participants from Kality Federal Prison and Kilinto Detention Center in Addis Ababa, as well as at Ziway Prison, which is located ∼170 KM south of Addis Ababa. Kality Federal Prison houses approximately 4,500 incarcerated individuals, while Ziway Prison houses 2,742 persons and accepts overflow transfers from Kality Prison. Kilinto Detention Center is situated near Kality Federal Prison and housed approximately 900 individuals. Healthcare services are provided by Kality Prison Hospital and the Ziway Prison Health Center, each equipped with four-module GeneXpert machines (Cepheid, Sunnyvale, CA, USA) for rapid TB detection. Incarcerated individuals from Kilinto utilize laboratory facilities at Kality. At the study sites, TB diagnosis relied mainly on passive case finding, with symptom-based screening performed at entry. All confirmed TB cases were isolated and received anti-TB treatment according to national guidelines.

### Study Population and Participant Enrollment Study Population

The study population consisted of incarcerated individuals from the study prisons and a detention center. Inclusion criteria were: age ≥18 years, ability and willingness to provide written informed consent, presence of cough of any duration with or without other TB-related symptoms, and ability to provide the required biological samples (sputum and oral swab) for laboratory analysis. Incarcerated individuals who had been on anti-TB treatment for more than two days were excluded.

### Participant Enrollment

Participant enrollment involved active screening, contact tracing, and inclusion of incarcerated individuals who visited prison clinics for TB evaluation. Of over 8,142 incarcerated individuals across the three study sites, 858 were screened between January 15 and October 31, 2025. The majority of screening activities were conducted at Kality Federal Prison. At this site, two trained prison nurses conducted monthly cell-to-cell visits in randomly selected prison cells throughout the study period. During these visits, resident inmates were screened for cough and other TB-related symptoms using a standardized screening protocol. Each month, a new set of cells was randomly selected, and screening was conducted over two consecutive days. In addition, incarcerated individuals identified as close contacts of confirmed TB cases were screened throughout the study period, and those who visited the prison clinic with TB-related symptoms between monthly screening cycles were also assessed for study eligibility.

Screening at Ziway Prison and Kilito Detention Center followed a similar approach; however, active screening was conducted only twice during the study period (May 5–7 and September 8–10, 2025). Contact tracing and assessment of incarcerated individuals who sought care passively were conducted continuously throughout the study period.

Following screening, incarcerated individuals meeting the inclusion criteria were enrolled in the study. Data were collected on socio-demographic and health characteristics, including age, sex, duration of imprisonment, history of sharing a cell with previously diagnosed TB patients, TB symptoms, past TB history, and HIV status. HIV status was obtained by self-report from participants; no HIV testing was performed as part of the study.

Screening was limited to resident incarcerated men living in the prisons. Entry screening of newly admitted individuals was not performed due to logistical constraints, and incarcerated women were not included. At Kality Federal Prison, women resided in a separate compound approximately 10 km from the men’s compound, and simultaneous screening of both compounds was not possible.

### Sample Collection and Analysis Sample Collection

Trained prison-based laboratory personnel collected the required samples from all participants. Two oral swabs were collected from each participant: an early-morning swab, followed by a spot swab collected 30 minutes later, both before eating, drinking, or tooth brushing. Each oral swab was collected by brushing the mid-dorsum of the tongue 10 times using FLOQSwabs (Tianzhu Sun Time Biotechnology, China). Swab heads were placed into a Falcon tube containing 234 µL of sterile 1× Tris-EDTA-Tween (TET) buffer and stored at 2–8 °C until processing within five days. Evidence shows that processing two swabs together, instead of a single swab, can improve the sensitivity of TB detection (17). The 15–30-minute interval between swab collections allows replenishment of Mtb on the tongue surface, ensuring independent and adequately loaded samples for testing (25).

Two consecutive morning sputum samples, each of sufficient volume (>3 mL), were collected. The first sample was tested immediately using Ultra, while the second was stored at –20 °C in the prison laboratory for 4-8 weeks before being transported in a cold box to the Ethiopian Public Health Institute (EPHI) in Addis Ababa for culture processing.

### Sample Analysis Sputum Xpert Ultra

Ultra testing was performed on the first morning sputum sample according to the manufacturer’s instructions, using a 2:1 ratio of SR to sample. The system processes samples internally within the cartridge and performs a hemi-nested qPCR targeting the *IS6110* and *IS1081* sequences, as well as the *rpoB* gene for rifampin resistance (23).

### Mycobacteria Growth Indicator Tube (MGIT) Culture

MGIT 960 (Becton Dickinson, Franklin Lakes, NJ, USA) liquid culture was performed at the National Tuberculosis Research and Reference Laboratory, EPHI, on the second sputum sample according to the manufacturer’s instructions (26). Sputum samples were decontaminated using the NALC–NaOH method (0.25% N-acetyl-L-cysteine, 1% NaOH), and the resulting sediments were inoculated into MGIT 960 tubes. Positive cultures were checked for contamination on Brain Heart Infusion (BHI) agar and examined for cord formation using Ziehl–Neelsen (ZN)–stained microscopy. Further identification was performed using the Capilia TB-Neo rapid test to confirm whether the isolate belonged to the Mtb complex (MTBC) or was non-MTBC (27). Laboratory personnel conducting culture testing were blinded to the oral swab Ultra results.

### Oral Swab Xpert Ultra

Oral swab Ultra testing was performed using a 2:1 ratio SR to sample. For each swab type, 466 µL of SR was added to 234 µL of TET buffer containing the swabs, giving a total volume of 700 µL. This analytically validated protocol, which yielded a final SR concentration of 16.6%, has previously been shown to have a lower limit of detection (LoD) of 30.3 colony-forming units (CFU) per 700 µL, substantially outperforming earlier protocols that used a final SR concentration of 66.6% (800 µL Tris–EDTA + 1,600 µL SR) and demonstrated a higher LoD of 75.5 CFU per swab (17, 24). The mixture was incubated at room temperature for 10 minutes. Subsequently, 500 µL of the suspension was transferred into an Xpert Ultra cartridge prefilled with 1.5 mL of TET buffer (final volume, 2 mL) and tested at the prison laboratory.

### Definitions

A composite microbiological reference standard (MRS) consisting of sputum Ultra and MGIT culture was used. Microbiologically confirmed TB was defined as detection of Mtb by MGIT culture and/or a positive sputum Ultra result at any semiquantitative level (high, medium, low, very low, or trace). All patients diagnosed with TB were promptly linked to appropriate treatment and care.

### Analysis

A REDCap database was used for data entry, and SPSS version 23 was used for data analysis (28). The sensitivity and specificity of oral swab Ultra were calculated using sputum Ultra and the MRS and are reported with 95% confidence intervals (CIs). Positive predictive value (PPV) and negative predictive value (NPV) were also estimated with corresponding 95% CIs. In addition, sensitivity of oral swab Xpert Ultra was calculated for those with sputum Xpert Ultra reference test across different bacillary loads. Differences in median threshold cycle (Ct) values between sputum Xpert Ultra and oral swab Xpert Ultra were assessed using the Wilcoxon signed-rank test.

## RESULTS

### Participant enrolment and diagnostic outcomes

Between January 15 and October 31, 2025, a total of 858 participants were screened for TB symptoms, of whom 240 were eligible for the study. In total, ten monthly screening cycles were conducted at Kality Federal Prison, whereas two screening cycles were performed at Ziway Prison and Kilito Detention Center. Figure 1 shows the participant enrollment and laboratory testing procedures. Out of 240 eligible participants, 221 were included in the final analysis. Nineteen participants were excluded from the analysis: 9 had contaminated MGIT cultures, 4 produced indeterminate results on oral swab Ultra, and 6 had indeterminate results on sputum Ultra. None of the indeterminate oral swab or sputum Ultra results were culture-positive, and the contaminated samples were also negative with sputum Ultra. A total of 34 participants (15.4%) were diagnosed with pulmonary TB based on MRS results. Among the 34 TB cases, 33 were positive by sputum Xpert Ultra and 33 by liquid culture, with one Ultra-positive/culture-negative and one Ultra-negative/culture-positive result. In addition, one nontuberculous mycobacterium (NTM) isolate was detected using culture.

**Figure 1:**
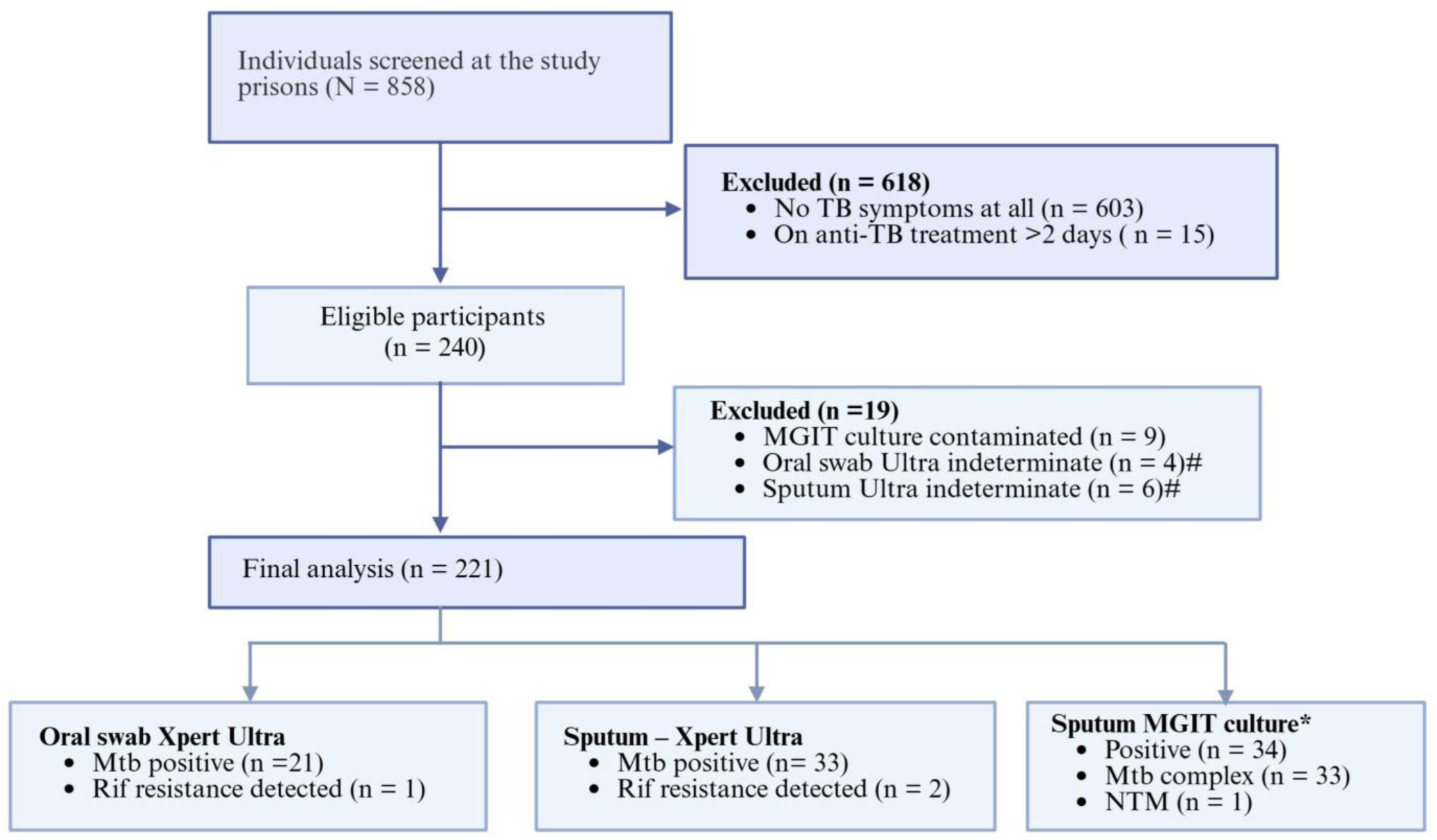
Participant enrollment and laboratory testing flow diagram. MGIT = Mycobacterium Growth Indicator Tube; Mtb = Mycobacterium tuberculosis; NTM = Nontuberculous mycobacteria; Rif = Rifampicin; TB = Tuberculosis. *Sputum Xpert+ → culture negative (n = I) and Sputum Xpert- → culture positive (n = 1); # indeterminate for Mtb detection

### Participants characteristics

Demographic and clinical characteristics of the participants are presented in Table 1. The majority of participants, 70.6% (156/221), were enrolled from Kality Federal Prison, and most (63.3%) were recruited through the monthly active screening approach. All participants were male, with a mean age of 30.5 years (SD ± 9.4) and a median body mass index (BMI) of 22.1 kg/m² (IQR: 20.4–23.8); 9% were underweight. A small proportion reported a previous TB diagnosis (2.7%) or were with HIV (3.6%). Nearly one-quarter of participants (23.9%) reported sharing a cell with a person later diagnosed with TB for at least one week. All participants reported having a cough; the prevalence of other symptoms was as follows: fever (38.0%), night sweats (32.6%), weight loss (24.4%), and hemoptysis (2.7%).

**Table 1.**
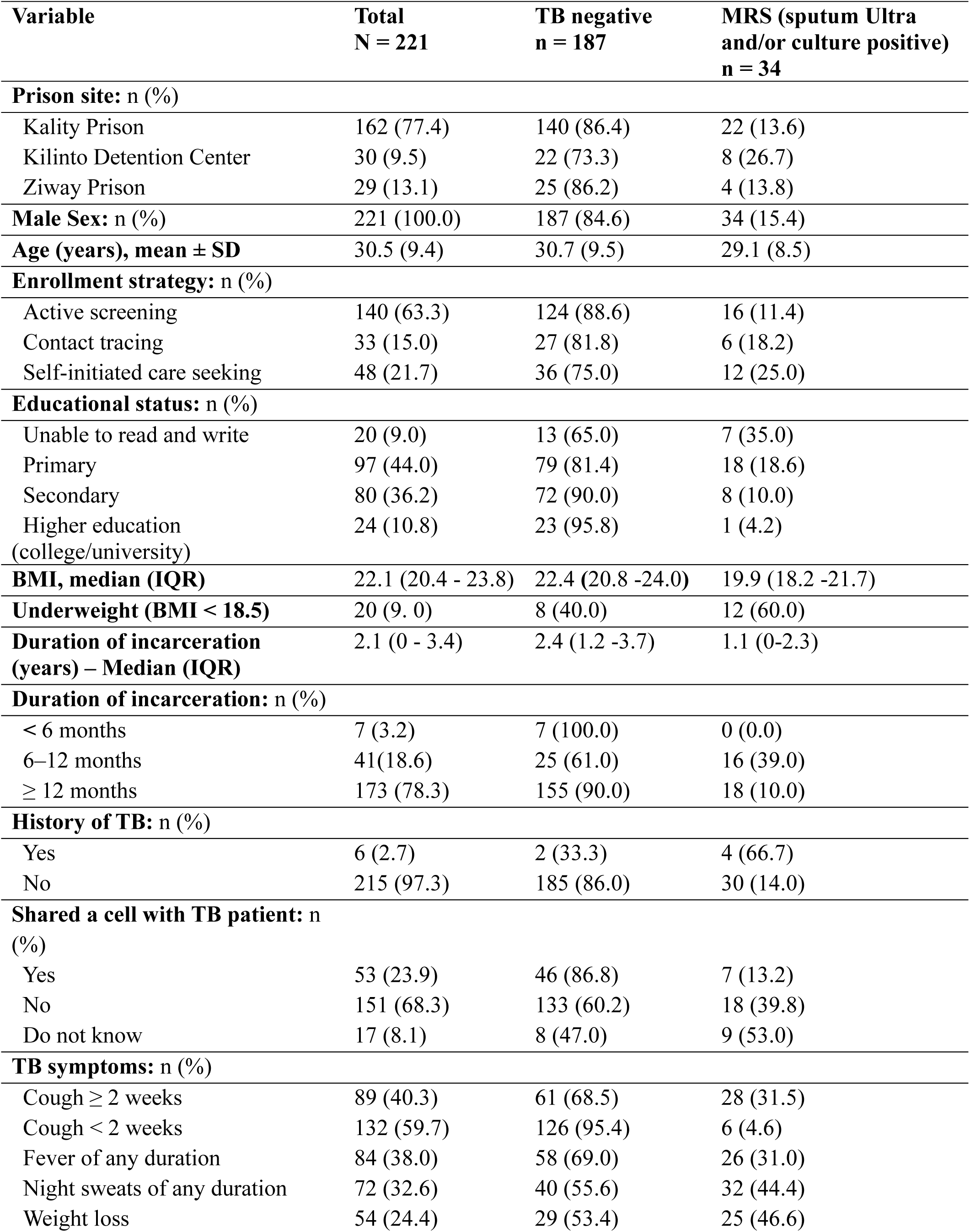
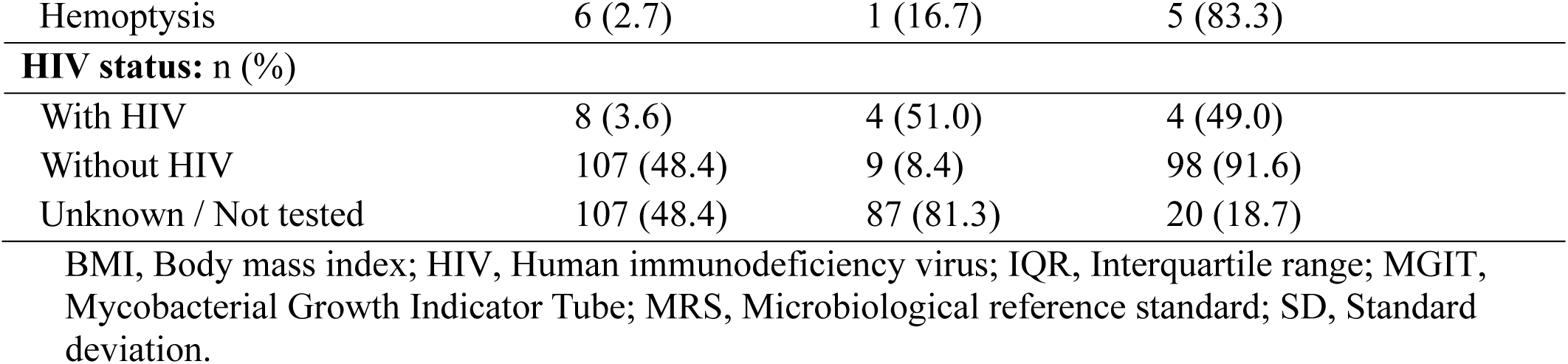
Demographic and clinical characteristics of participants in oral swab ultra compared with sputum-based microbiological reference standard.

| **Variable** | **Total**<br>**N = 221** | **TB negative**<br>**n = 187** | **MRS (sputum Ultra**<br>**and/or culture positive)**<br>**n = 34** |
| --- | --- | --- | --- |
| **Prison site: n (%)** |  |  |  |
| Kality Prison | 162 (77.4) | 140 (86.4) | 22 (13.6) |
| Kilinto Detention Center | 30 (9.5) | 22 (73.3) | 8 (26.7) |
| Ziway Prison | 29 (13.1) | 25 (86.2) | 4 (13.8) |
| **Male Sex: n (%)** | 221 (100.0) | 187 (84.6) | 34 (15.4) |
| **Age (years), mean ± SD** | 30.5 (9.4) | 30.7 (9.5) | 29.1 (8.5) |
| **Enrollment strategy: n (%)** |  |  |  |
| Active screening | 140 (63.3) | 124 (88.6) | 16 (11.4) |
| Contact tracing | 33 (15.0) | 27 (81.8) | 6 (18.2) |
| Self-initiated care seeking | 48 (21.7) | 36 (75.0) | 12 (25.0) |
| **Educational status: n (%)** |  |  |  |
| Unable to read and write | 20 (9.0) | 13 (65.0) | 7 (35.0) |
| Primary | 97 (44.0) | 79 (81.4) | 18 (18.6) |
| Secondary | 80 (36.2) | 72 (90.0) | 8 (10.0) |
| Higher education<br>(college/university) | 24 (10.8) | 23 (95.8) | 1 (4.2) |
| **BMI, median (IQR)** | 22.1 (20.4 - 23.8) | 22.4 (20.8 -24.0) | 19.9 (18.2 -21.7) |
| **Underweight (BMI < 18.5)** | 20 (9.0) | 8 (40.0) | 12 (60.0) |
| **Duration of incarceration**<br>**(years) – Median (IQR)** | 2.1 (0 - 3.4) | 2.4 (1.2 -3.7) | 1.1 (0-2.3) |
| **Duration of incarceration: n (%)** |  |  |  |
| < 6 months | 7 (3.2) | 7 (100.0) | 0 (0.0) |
| 6–12 months | 41 (18.6) | 25 (61.0) | 16 (39.0) |
| ≥ 12 months | 173 (78.3) | 155 (90.0) | 18 (10.0) |
| **History of TB: n (%)** |  |  |  |
| Yes | 6 (2.7) | 2 (33.3) | 4 (66.7) |
| No | 215 (97.3) | 185 (86.0) | 30 (14.0) |
| **Shared a cell with TB patient: n (%)** |  |  |  |
| Yes | 53 (23.9) | 46 (86.8) | 7 (13.2) |
| No | 151 (68.3) | 133 (60.2) | 18 (39.8) |
| Do not know | 17 (8.1) | 8 (47.0) | 9 (53.0) |
| **TB symptoms: n (%)** |  |  |  |
| Cough ≥ 2 weeks | 89 (40.3) | 61 (68.5) | 28 (31.5) |
| Cough < 2 weeks | 132 (59.7) | 126 (95.4) | 6 (4.6) |
| Fever of any duration | 84 (38.0) | 58 (69.0) | 26 (31.0) |
| Night sweats of any duration | 72 (32.6) | 40 (55.6) | 32 (44.4) |
| Weight loss | 54 (24.4) | 29 (53.4) | 25 (46.6) |
| Hemoptysis | 6 (2.7) | 1 (16.7) | 5 (83.3) |
| **HIV status: n (%)** |  |  |  |
| With HIV | 8 (3.6) | 4 (51.0) | 4 (49.0) |
| Without HIV | 107 (48.4) | 9 (8.4) | 98 (91.6) |
| Unknown / Not tested | 107 (48.4) | 87 (81.3) | 20 (18.7) |

BMI, Body mass index; HIV, Human immunodeficiency virus; IQR, Interquartile range; MGIT, Mycobacterial Growth Indicator Tube; MRS, Microbiological reference standard; SD, Standard deviation.

### Oral Swab Xpert Ultra Diagnostic Accuracy

Relative to sputum Ultra, oral swab Ultra demonstrated a sensitivity of 63.6% (21/33; 95% CI: 45.1–79.6%) and a specificity of 100%. Against the MRS, which included both sputum Ultra and MGIT culture, the sensitivity was 61.8% (21/34; 95% CI: 44.7–76.6%), with specificity remaining 100%. Notably, sensitivity improved to 80.8% (21/26; 95% CI: 60.6–93.4%) when samples with “trace” sputum results were excluded from the analysis.

### Diagnostic Accuracy by Semiquantitative Grade

The sensitivity of oral swab Ultra increased with higher sputum bacillary loads, reaching 100% among patients with medium to high bacillary loads with sputum Ultra. Semi-quantitative results were generally lower in oral swabs than in sputum samples. None of the seven sputum ‘trace’ results were detected by oral swabs. The mean number of TB symptoms was higher among patients with high and medium bacillary loads (Table 3). Overall, patients diagnosed with TB had an average of 3.9 TB-related symptoms (SD ± 0.8).

**Table 2:** Sensitivity and specificity of oral swab ultra compared with sputum-based microbiological reference standard.

| Parameter | Sputum Xpert Ultra reference standard<br>n/N, % (95% CI) | Microbiologic reference standard<br>n/N, % (95% CI) |
| --- | --- | --- |
| Sensitivity | 21/33, 63.6 (45.1-79.6) | 21/34, 61.8 (44.7-76.6) |
| Specificity | 188/188, 100 (98.1-100) | 187/187, 100 (97.9-100) |
| PPV | 21/21, 100 (83.9-100) | 21/21, 100 (83.9-100) |
| NPV | 188/200, 94 (89.7-96.8) | 187/200, 93.5 (89.1-96.3) |

CI, confidence interval; PPV, positive predictive value; NPV, negative predictive value.

**Table 3:** Diagnostic accuracy of oral swab ultra compared to sputum-based microbiological reference standard.

| **Sputum Xpert**<br>**Ultra grade** | **Detected by oral swabs**<br>**% (n/N)** | **Mean number of symptoms<sup>a</sup>**<br>**(±SD)** |
| --- | --- | --- |
| High | 100 (4/4) | 4.1 (0.7) |
| Medium | 100 (5/5) | 4.0 (0.8) |
| Low | 81 (9 /11) | 3.8 (0.8) |
| Very low | 50 (3/6) | 3.8 (0.8) |
| Trace | 0 (0/7) | 3.8 (0.8) |
| **All** | **63.6 (21/33)** | **3.9 (0.8)** |

a, cough, fever, night sweats, weight loss, hemoptysis; SD, Standard deviation.

The distribution of Ultra Threshold-cycle (Ct) values for sputum and oral swab specimens is shown in Figure 2. Median Ct values for *IS6110/IS1081* were 24.2 (IQR 20.0–28.5) for oral swab Ultra and 20.2 (IQR 16.6–23.8) for sputum Ultra. Ct values were generally higher for oral swab Ultra than for sputum Ultra, with a median difference of 4.0 Ct cycles (95% CI: 1.6–6.4); however, this difference was not statistically significant (Wilcoxon signed-rank test, p = 0.29).

**Figure 2:**
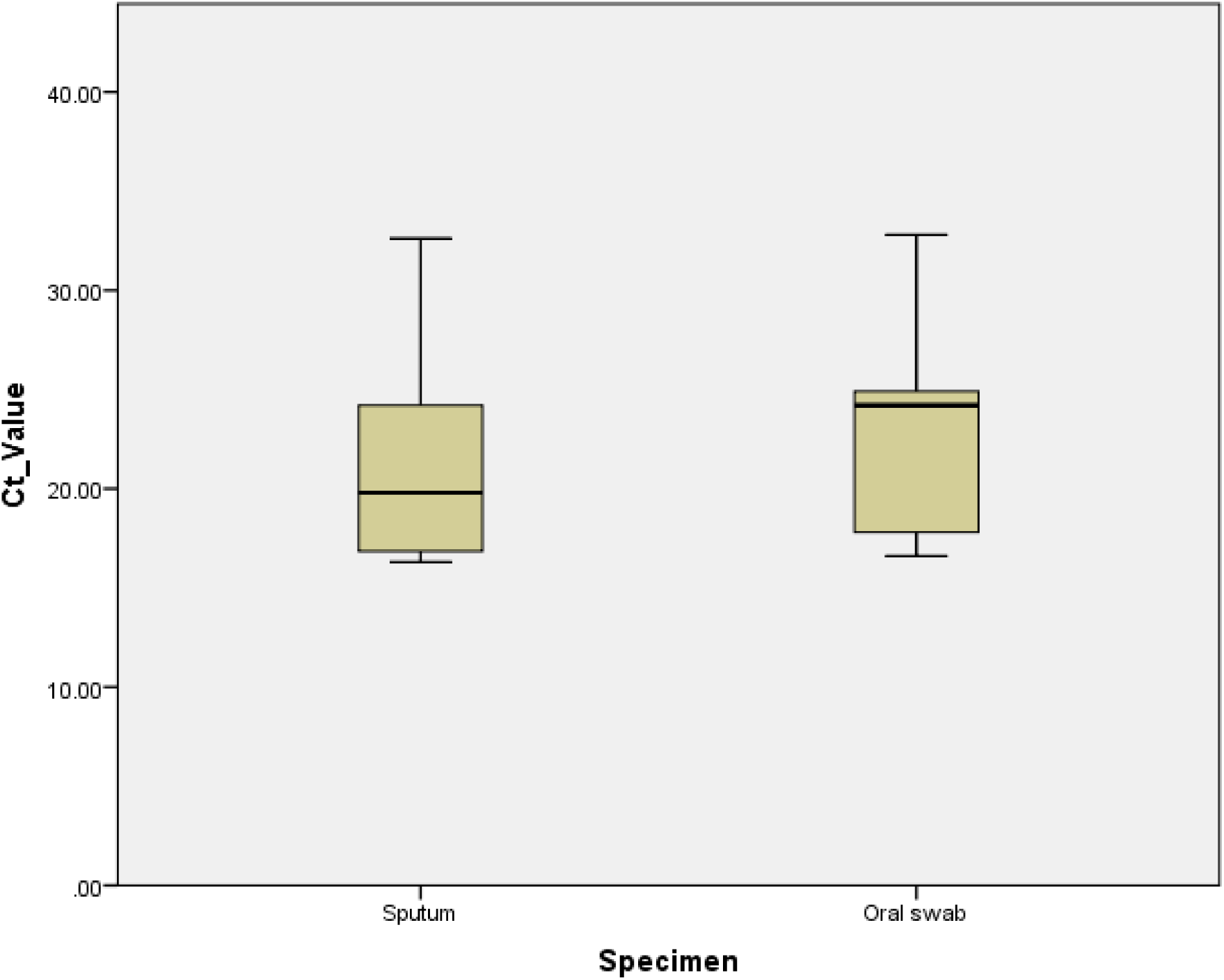
Distribution of Xpert Ultra Ct values for sputum and oral swab specimens

## DISCUSSION

We demonstrate the performance and operational feasibility of oral swab Ultra testing for enhancing TB detection for the first time in Ethiopian prison settings. In the context of the high TB prevalence observed in this population, oral swab sampling with Ultra testing showed sensitivities of 63.6% versus sputum Xpert Ultra and 61.8% versus MRS, with 100% specificity in both comparisons. Notably, oral Ultra testing was positive in all participants with medium or high sputum bacillary loads, showing that mycobacterial burden influences oral swab performance. Importantly, OSA detected 70.6% of patients with low or very low bacillary loads, suggesting utility even in paucibacillary disease. These findings support the integration of sputum-free testing methods in prisons, which could enable earlier case detection, timely treatment, and reduced transmission within high-risk settings.

In our study, OSA did not detect TB cases that had ‘trace’ bacilli, as has been reported previously (17, 29). However, these ‘trace’ positive patients may also contribute to the spread of TB within the prison setting, especially as all but one of such cases in our study were culture positive. A study has shown that patients who are ‘trace’ positive on sputum samples screened for TB had a significant risk of developing symptomatic disease within two years (30). Further research should focus on optimizing oral swab collection and processing methods to detect ‘trace’ TB cases (31).

Previous studies on oral swab Ultra were mainly conducted in non-prison healthcare facility settings, making direct comparisons difficult, as prisons represent distinct environments for TB (4). The sensitivity in our study (61.8% vs. MRS) was slightly lower than that reported in the Ugandan study (72.4%) (17) and studies from South Africa (59.0-78.1%) (18, 19, 22). The discrepancies could be partly attributed to differences in the recruitment approach for participants. The aforementioned studies recruited participants who were seeking healthcare services and had a cough for two weeks or more, which tends to indicate a high bacillary load. However, in our study, we recruited participants with a cough of any duration, which could have increased the proportion of cases with lower bacillary loads, where the oral swab’s sensitivity is expected to be lower. This is reflected in our findings, where 45.5% (15/33) of TB cases had trace to very low bacillary loads, compared with proportions ranging from 14.3% to 21.2% in the comparative studies (17–19). However, the 61.8% sensitivity observed in this study was higher than that reported in other health facility–based studies, including a study conducted in Lima, Peru, which reported a sensitivity of 45% (20).

The implication of oral swabs extends beyond test accuracy because their use may be beneficial in improving diagnostic yield, as oral swabs are easier, safer, and less stigmatizing than sputum collection (32). These advantages contribute to increasing the participation in the TB screening and hence potentially offsetting their relatively low sensitivity. A recent study showed that the yield of the tongue swab (also known as oral swab) test using the MiniDock MTB was similar to that of the sputum test (3.8% vs. 4.1%) (33). In our study, all eligible inmates provided an oral swab, and 19.0% of TB cases had only mild symptoms, reinforcing their potential role in detecting low-bacillary-burden TB and potentially implying an increase in diagnostic yield.

Compared with a prison-based study from Brazil, which reported a sensitivity of 51% using oral swab Ultra (21), our study achieved higher sensitivity. This improvement may be explained by the methodological innovations not widely tested in prison settings. We used a dual-swab approach, including both early-morning and spot swabs, with a lower final SR concentration in oral swab processing, which could have enhanced the recovery of low-level mycobacterial DNA and improved our test performance (34). Differences in study population characteristics, TB burden, and disease stage may also have contributed to the observed variation in sensitivity.

Several studies have evaluated SR-based oral swab processing in comparison with heat-based methods for TB detection using Ultra. Andama et al. reported LOD of 76.5 CFU/swab using a double-swab SR method (final SR 66.6%) in an analytical study, which was reduced to 22.3 CFU/swab with boiling-based inactivation, although boiling caused over-pressurization errors (17). More recently, Chilambi et al compared four swab processing methods, finding the lowest LOD with 1:1 diluted SR (22.7 CFU/700 µL), followed by 2:1 diluted SR (30.3 CFU/700 µL) and neat SR (30.9 CFU/700 µL), while heat-based inactivation yielded the highest LOD (77.6 CFU/700 µL) (24). Although the 1:1 dilution achieved the lowest LOD, the 2:1 diluted SR protocol provided a better balance between detection sensitivity, biosafety, and sample stability. These analytical findings suggest that optimized SR dilution enhances mycobacterial DNA recovery, which may translate into improved clinical diagnostic performance. Our clinical validation study adopted this 2:1 diluted SR protocol (30.3 CFU/700 µL) using double swabs and demonstrated promising performance for *Mtb* detection in prison settings.

Our results also demonstrate the operational feasibility of the oral swab Ultra in prisons. TB control in SSA prisons, such as in Ethiopia, is hindered by several issues, such as lack of space to isolate symptomatic incarcerated individuals and poor infection control practices, and TB-related stigma associated with the provision of sputum samples (4, 35). TB prevention and treatment in prisons is further complicated by the inability of many inmates to produce sputum; for example, in a Brazilian mass screening study, about two-thirds of inmates were unable to provide sputum samples, emphasizing the need for non-sputum–based diagnostic approaches such as oral swabs (36).

Although comparisons with previous prison-based prevalence studies in Ethiopia are limited due to differences in screening approaches, our study—which was primarily designed to assess diagnostic accuracy—found a high TB prevalence of 15.4% among symptomatic participants (5, 37). The detection of two rifampicin-resistant cases represents a serious public health concern. Prevalence peaked in those incarcerated for 6–12 months, apparently representing the development of undiagnosed community-acquired TB and early prison-acquired transmission. However, 10% prevalence among individuals incarcerated for more than 12 months also indicates that symptomatic cases had remained undiagnosed within the prison for prolonged periods, suggesting ongoing in-prison transmission of TB. Nearly one-quarter of participants reported sharing a cell for at least one week with a person later diagnosed with TB, and 13.2% of these individuals subsequently developed TB, further suggesting the ongoing transmission within the prisons. Overall, these findings highlight gaps in the routine passive case-finding approach, even in the presence of GeneXpert machines in the prisons, and emphasize the need to prioritize systematic active case-finding strategies for both new entrants and residents to ensure early detection and treatment of TB cases. Adding oral swab Ultra into regular TB control programs in these prisons could further enhance case detection by identifying individuals with subclinical or asymptomatic TB, thereby facilitating earlier treatment and reducing disease spread.

A key strength of our study is the use of an analytically validated protocol employing dual oral swabs, including early-morning oral swabs. We validated the protocol in real-world, high-risk prisons in an SSA region using a robust MRS. By including individuals with coughs of any duration, we were able to assess the performance of oral swabs in paucibacillary TB, a group that is often underrepresented or missed in facility-based studies.

Limitations include the inability to fully attribute improved performance to the optimized protocol, as we did not conduct head-to-head comparisons with alternative oral swab techniques. Screening was limited to male incarcerated individuals due to logistical constraints. Oral swab performance is not expected to differ by sex, but this limits the generalizability of the findings to the entire prison population. Despite these, the study provides important pragmatic data on the diagnostic accuracy and feasibility of an optimized oral swab Ultra in a high-burned prison setting, in an area where data are scarce.

## Conclusion

In conclusion, a customized, analytically validated SR-based method for oral swab processing and testing on the Ultra showed moderate to high sensitivity in high-burden prison settings in SSA, with performance largely dependent on sputum bacillary load. It has been shown to be acceptable by the participants and could be used as a complementary tool in TB mass screening initiatives in Ethiopian prison settings and other similar settings in SSA. However, its inability to detect TB cases with trace bacillary loads suggests the need for further research for optimization, particularly for asymptomatic or paucibacillary patients.

## Data Availability

All data produced in the present work are contained in the manuscript.

## Acknowledgments

This work was supported by the National Institutes of Health (NIH) Fogarty International Center Global Infectious Diseases Grant (D43TW009127) through the Ethiopia–Emory TB Research Training Program (EETB-RTP) and the NIH National Institute of Allergy and Infectious Diseases [K24AI190403, P30AI168386], NIH TRAC grant, and NIH K24 mentoring grant.

We are grateful to EPHI for providing facilities for sputum culture and the swabs, to the Armauer Hansen Research Institute (AHRI), Ethiopia, for providing TET buffer, and to the prison health professionals at the study sites and the laboratory staff at EPHI for their valuable support.

The funders of the study had no role in study design, data analysis, data interpretation, or writing of the report.

## REFERENCES

1. World Health Organization. Global tuberculosis report. 2025.

2. Placeres AF, de Almeida Soares D, Delpino FM, Moura HSD, Scholze AR, Dos Santos MS, et al. Epidemiology of TB in prisoners: a metanalysis of the prevalence of active and latent TB. BMC Infectious Diseases. 2023;23(1):20.

3. Baussano I, Williams BG, Nunn P, Beggiato M, Fedeli U, Scano F. Tuberculosis incidence in prisons: a systematic review. PLoS medicine. 2010;7(12):e1000381.

4. Mera HB, Wagnew F, Akelew Y, Hibstu Z, Berihun S, Tamir W, et al. Prevalence and predictors of pulmonary tuberculosis among prison inmates in Sub-Saharan Africa: a systematic review and meta-analysis. Tuberculosis Research and Treatment. 2023;2023(1):6226200.

5. Genet A, Girma A. Magnitude, associated risk factors, and trend comparisons of identified tuberculosis types among prisons in Ethiopia: A systematic review and meta-analysis. Health science reports. 2024;7(1):e1789.

6. Miyahara R, Piboonsiri P, Chiyasirinroje B, Imsanguan W, Nedsuwan S, Yanai H, et al. Risk for Prison-to-Community Tuberculosis Transmission, Thailand, 2017–2020. Emerging Infectious Diseases. 2023;29(3):477–83.

7. Walter KS, Dos Santos PCP, Gonçalves TO, da Silva BO, da Silva Santos A, de Cássia Leite A, et al. The role of prisons in disseminating tuberculosis in Brazil: A genomic epidemiology study. The Lancet Regional Health–Americas. 2022;9.

8. Pai M, Dewan PK, Swaminathan S. Transforming tuberculosis diagnosis. Nat Microbiol. 2023:1–4.

9. Adane K, Spigt M, Ferede S, Asmelash T, Abebe M, Dinant G-J. Half of pulmonary tuberculosis cases were left undiagnosed in prisons of the Tigray region of Ethiopia: implications for tuberculosis control. PloS one 2016;11(2):e0149453.

10. Nyasulu PS, Hui DS, Mwaba P, Tamuzi JL, Sakala DY, Ntoumi F, et al. Global perspectives on tuberculosis in prisons and incarceration centers-Risk factors, priority needs, challenges for control and the way forward. IJID regions. 2025;14:100621.

11. Papadopoulou P, Gaeddert M, Gupta-Wright A, Denkinger C, Marx F. Sputum availability and quality in country-level TB prevalence surveys. IJTLD open. 2024;1(11):528–30.

12. Wang Z, Li H, Song S, Sun H, Dai X, Chen M, et al. Transmission of tuberculosis in an incarcerated population during the subclinical period: A cross-sectional study in Qingdao, China. Frontiers in Public Health. 2023;11:1098519.

13. Mangu CD, Clowes P, van den Hombergh J, Mwakabenga C, Mwanyonga S, Ambindwile J, et al. New Admissions and Asymptomatic TB Cases fuel TB Epidemic in Prisons, a Cross Sectional Survey in Tanzania. BMJ Yale 2023:2023.08. 23.23294476.

14. Frascella B, Richards AS, Sossen B, Emery JC, Odone A, Law I, et al. Subclinical tuberculosis disease—a review and analysis of prevalence surveys to inform definitions, burden, associations, and screening methodology. Clinical Infectious Diseases. 2021;73(3):e830–e41.

15. World Health Organization. WHO operational handbook on tuberculosis. Module 2: screening - systematic screening for tuberculosis disease. . 2021.

16. Byrne RL, Wingfield T, Adams ER, Banu S, Bimba JS, Codlin A, et al. Finding the missed millions: innovations to bring tuberculosis diagnosis closer to key populations. BMC Global and Public Health. 2024;2(1):33.

17. Andama A, Whitman GR, Crowder R, Reza TF, Jaganath D, Mulondo J, et al. Accuracy of tongue swab testing using Xpert MTB-RIF Ultra for tuberculosis diagnosis. J Clin Microbiol. 2022;60(7):e00421–22.

18. David A, Singh L, Peloakgosi-Shikwambani K, Nsingwane Z, Molepo V, Cangelosi G, et al. Diagnostic accuracy of self-collected tongue swabs for Mycobacterium tuberculosis complex detection in individuals being assessed for tuberculosis in South Africa using the Xpert MTB/RIF Ultra assay. Clinical Microbiology and Infection. 2025.

19. Wood RC, Luabeya AK, Dragovich RB, Olson AM, Lochner KA, Weigel KM, et al. Diagnostic accuracy of tongue swab testing on two automated tuberculosis diagnostic platforms, Cepheid Xpert MTB/RIF Ultra and Molbio Truenat MTB Ultima. Journal of Clinical Microbiology. 2024;62(4):e00019–24.

20. Mesman AW, Calderon R, Soto M, Coit J, Aliaga J, Mendoza M, et al. Mycobacterium tuberculosis detection from oral swabs with Xpert MTB/RIF ULTRA: a pilot study. BMC research notes. 2019;12(1):349.

21. Lima F, Santos AS, Oliveira RD, Silva CC, Gonçalves CC, Andrews JR, et al. Oral swab testing by Xpert® MTB/RIF Ultra for mass tuberculosis screening in prisons. Journal of clinical tuberculosis and other mycobacterial diseases. 2020;19:100148.

22. Rockman L, Abdulgader S, Minnies S, Palmer Z, Naidoo CC, Naidoo D, et al. Oral washes and tongue swabs for Xpert MTB/RIF Ultra-based tuberculosis diagnosis in people with and without the ability to make sputum. Research Square. 2025:rs. 3. rs-6225530.

23. Chakravorty S, Simmons AM, Rowneki M, Parmar H, Cao Y, Ryan J, et al. The new Xpert MTB/RIF Ultra: improving detection of Mycobacterium tuberculosis and resistance to rifampin in an assay suitable for point-of-care testing. MBio. 2017;8(4):10.1128/mbio.00812-17.

24. Chilambi GS, Reiss R, Daivaa N, Banada P, De Vos M, Penn-Nicholson A, et al. Optimal processing of tongue swab samples for Mycobacterium tuberculosis detection by the Xpert MTB/RIF Ultra assay. Microbiology Spectrum. 2025;13(3):e02403–24.

25. Wood RC, Luabeya AK, Weigel KM, Wilbur AK, Jones-Engel L, Hatherill M, et al. Detection of Mycobacterium tuberculosis DNA on the oral mucosa of tuberculosis patients. Scientific reports. 2015;5(1):8668.

26. Siddiqi SH, Rüsch-Gerdes SJ, Foundation for innovative new Diagnostic : Borstel G. Mycobacteria Growth Indicator Tube Culture Drug Susceptibility Demonstration Project. 2006.

27. Muyoyeta M, Mwanza WC, Kasese N, Cheeba-Lengwe M, Moyo M, Kaluba-Milimo D, et al. Sensitivity, specificity, and reproducibility of the Capilia TB-Neo assay. Journal of clinical microbiology. 2013;51(12):4237–9.

28. Harris PA, Taylor R, Minor BL, Elliott V, Fernandez M, O’Neal L, et al. The REDCap consortium: building an international community of software platform partners. Journal of biomedical informatics. 2019;95:103208.

29. Ajide B, Moe CA, Barrameda J, Chirwa M, Rockman L, de Haas P, et al. Tongue swab xpert MTB/RIF ultra testing for tuberculosis using a revised consensus protocol: a multi-country diagnostic accuracy study. medRxiv. 2025.

30. Sung J, Nantale M, Nalutaaya A, Biché P, Mukiibi J, Akampurira J, et al. Long-term risk of tuberculosis among individuals with Xpert Ultra trace screening results in Uganda: a longitudinal follow-up study. The Lancet Infectious Diseases. 2025.

31. Zhang F, Wang Y, Zhang X, Liu K, Shang Y, Wang W, et al. Diagnostic accuracy of oral swab for detection of pulmonary tuberculosis: a systematic review and meta-analysis. Frontiers in medicine. 2024;10:1278716.

32. Luabeya AK, Wood RC, Shenje J, Filander E, Ontong C, Mabwe S, et al. Noninvasive detection of tuberculosis by oral swab analysis. Journal of clinical microbiology. 2019;57(3):10.1128/jcm.01847-18.

33. Moe CA, Luswata RK, Barrameda AJ, Le H, Muzazu S, Crowder R, et al. Diagnostic yield of tongue swab-compared to sputum-based molecular testing for tuberculosis in four high-burden countries. medRxiv. 2025:2025.07. 03.25330836.

34. Wang Y, Ma Z, Liu Z, Dong X, Shu W, Wei M, et al. Tongue swab-based molecular diagnostics for pulmonary tuberculosis and drug resistance in adults: A prospective multicenter diagnostic accuracy study. Journal of Infection. 2025:106517.

35. Adane K, Spigt M, Ferede S, Asmelash T, Abebe M, Dinant G-J. Half of pulmonary tuberculosis cases were left undiagnosed in prisons of the Tigray region of Ethiopia: implications for tuberculosis control. PLoS One. 2016;11(2):e0149453.

36. Paião DSG, Lemos EF, Carbone AdSS, Sgarbi RVE, Junior AL, da Silva FM, et al. Impact of mass-screening on tuberculosis incidence in a prospective cohort of Brazilian prisoners. BMC infectious diseases. 2016;16(1):533.

37. Adane K, Spigt M, Ferede S, Asmelash T, Abebe M, Dinant G-JJPo. Half of pulmonary tuberculosis cases were left undiagnosed in prisons of the Tigray region of Ethiopia: implications for tuberculosis control. 2016;11(2):e0149453.

